# Community pharmacy responses to online reviews: a national cross-sectional study

**DOI:** 10.64898/2026.11.21.26368650

**Authors:** Daho Samboré, Louise Meksem, Flora Huang, Julien Godet

## Abstract

**Objectives:** Online reviews constitute a source of information on the experience of users in community pharmacies. The way in which pharmacists engage with them remains poorly documented. The objective was to describe the frequency, determinants, and content of the responses provided by French pharmacy teams to reviews published on Google Maps.

**Methods:** Cross-sectional observational study. Public reviews from nearly all French community pharmacies (approximately 20 000) were collected automatically between September and October 2026. For each review, the rating, the text of the review, the time since its publication, and the text of the response, if it existed, were collected. Factors associated with the response were studied using logistic regression. Sentiment of the reviews and the tone of the responses were automatically coded according to a predefined grid. The presence of reviews generated by artificial intelligence, as well as potential interventions on the ratings, were explored.

**Results:** In total, 1 008 024 extractions (729 520 reviews and 278 504 responses) concerning 19 985 pharmacies were analysed. A response was present for 38% of reviews and 67.5 % of pharmacies had responded at least once. The response rate was higher for negative reviews (one or two stars) than for positive reviews (five stars) (adjusted OR 1.35; 95 % CI [1.33–1.38]; adjusted risk difference +3.6 percentage points; 95 % CI [3.3–3.8]), and these responses were about three times as long (adjusted geometric mean ratio 3.03; 95 % CI [3.01–3.06]). The tone of the responses was very widely judged to be appropriate. Comments generated by artificial intelligence seem to be appearing progressively and represent approximately 3-4% in 2026. The removal of poor ratings appears to be marginal.

**Conclusions:** Community pharmacists regularly and appropriately respond to online reviews, including, and in particular, less positive ones. These results could serve in the development of specific professional recommendations aimed at securing this practice.

## Introduction

Online review platforms play a growing role in the choice of a healthcare professional or establishment (1–4). Online reviews provide valuable information on patient experience, although they are unlikely to be suitable indicators of clinical quality or a sound basis for ranking healthcare professionals. (5). Among these platforms, Google Maps (Google LLC, Mountain View, CA, USA) allows users to associate a rating and freely written comments with each establishment. The community pharmacy, as a local business and a primary care setting, is not exempt from this public evaluation. With nearly 20 000 pharmacies in France (6), the pharmacy network constitutes one of the healthcare structure groups most exposed to online reviews. Several studies have used Google reviews to describe user experience in community pharmacies. In Saudi Arabia, an analysis of more than 17 000 reviews across 512 pharmacies shows that the propensity to write a review depends on the user’s perception (7). In Canada, a study of pharmacy reviews in British Columbia identifies professionalism and waiting time as the main themes expressed (8). In China, the themes addressed in comments on online pharmacies mainly concerned logistics, the price of medicines, customer service, and the effect of the medications (9). These studies focus on what users say, but they do not provide information on what pharmacists do with this information (10–13). Similarly, while European and French literature is beginning to take an interest in the digital transformation of health and the influence of review platforms on patient choice, research remains largely focused on the global perception of e-health and largely centered on content analysis of user comments, leaving the professionals’ uptake of this information largely unexplored (14–16). To date, precise and documented national data are lacking on how community pharmacists in France actively manage their e-reputation, particularly through the writing of responses to comments, thus leaving an analytical gap regarding the communication strategies adopted by professionals in response to user feedback. The healthcare professional’s response is an integral part of the exchange via these social platforms. Experimental data obtained for physicians indicate that the presence of a response mitigates the effect of negative reviews on users’ choice intentions (4). However, responding exposes the healthcare professional to a specific risk. Confirming that a person visited the establishment, or mentioning their care, may constitute a breach of professional secrecy, even when the user has made these elements public themselves (17). In France, the pharmacist is bound by professional secrecy and ethical rules governing their communication. The code of ethics for pharmacists was completely overhauled in March 2026 and recognises digital tools as information and communication supports while maintaining safeguards (18). The pharmacist may communicate by any means, including the internet, regarding their skills, their background, and their conditions of practice. This communication must be fair and honest, without third-party testimonials and without comparison with other pharmacists. Concepts of fair communication and professional dignity imply, for example, that the tone of responses, especially to negative reviews, affects the dignity of the profession. In this context, it is interesting to provide an overview of how pharmacy teams appropriate online reviews. To our knowledge, this work has not been described quantitatively and no national-scale study has been conducted.

The main objective of this work is to estimate the proportion of Google Maps reviews that received a response from the pharmacy in France. Secondary objectives are to identify the characteristics of the reviews and pharmacies associated with the existence of a response, to describe the tone and content of these responses, and to explore the appearance of comments generated by conversational tools as well as potential data manipulations.

## Materials and methods

### Study design and data

This was a cross-sectional, descriptive, and analytical observational study based on publicly accessible online data. The 20 003 community pharmacies were identified in the extraction of the open data National File of Health and Social Establishments (*Fichier National des Établissements Sanitaires et Sociaux* FINESS) published on data.gouv.fr (version dated 9 September 2026). Their Google Maps reviews were collected automatically between September and October 2026: rating (one to five stars), text, age of the review, and, where applicable, the text and age of the pharmacy team’s response, as well as the average rating of each profile were collected. Duplicates were removed (Supplementary Methods S1).

### Description of responses

The analyses focused on reviews containing a readable textual comment. A review was considered to have received a response when the latter contained at least one word. The response rate (Wilson 95 % CI) and the length of responses in number of words (median and interquartile range, IQR) were calculated for France as a whole and by region (Supplementary Methods S2.1).

### Primary endpoint

The primary endpoint was the presence of a response from the pharmacy team to a review. Reviews were classified as negative (one or two stars) or positive (five stars). The association between this polarity and the probability of response was estimated using a mixed-effects logistic regression, with a random intercept per pharmacy, adjusted for the length and age of the review, the pharmacy’s average rating, and its region. The results are expressed as adjusted odds ratios (OR, 95 % CI) and adjusted risk difference (percentage points). The length of the responses was compared between negative and positive reviews using a linear mixed model on the logarithm transform of the number of words, with the same adjustments, and expressed as an adjusted geometric mean ratio (aGMR) (Supplementary Methods S2.2).

### Nature and tone of responses

The review–response pairs were automatically coded by a large language model (LLM) run locally (Gemma 4 (19), 12 billion parameters; temperature 0; constrained JSON output), without data transmission to a third party. For each pair, the model coded the sentiment of the review, as well as the primary function of the response (a grid of thirteen inductively constructed categories, Table 1), its tone, the appropriateness of this tone for the review, and its degree of personalisation. The analysis of the response tone was conducted on a sample stratified by rating of 81 953 pairs; global estimates were weighted by the inverse of the sampling fractions (Supplementary Methods S3).

**Table 1.**
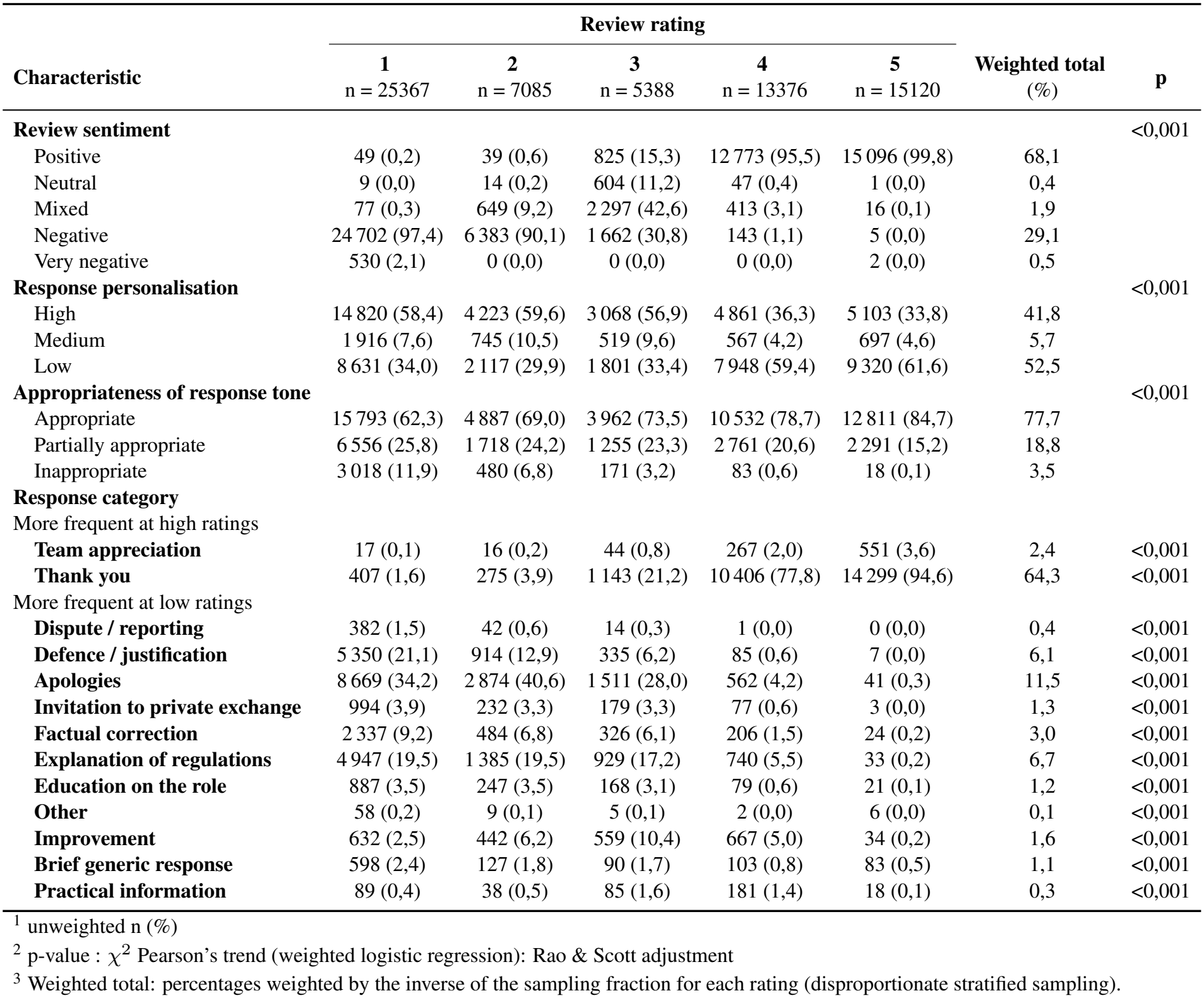
Characteristics of reviews and responses according to the rating assigned.

| **Characteristic** | **Review rating** |  |  |  |  | **Weighted total**<br>(%) | **p** |
|---|---|---|---|---|---|---|---|
|  | **1**<br>n = 25367 | **2**<br>n = 7085 | **3**<br>n = 5388 | **4**<br>n = 13376 | **5**<br>n = 15120 |  |  |
| **Review sentiment** |  |  |  |  |  |  | <0,001 |
| Positive | 49 (0,2) | 39 (0,6) | 825 (15,3) | 12 773 (95,5) | 15 096 (99,8) | 68,1 |  |
| Neutral | 9 (0,0) | 14 (0,2) | 604 (11,2) | 47 (0,4) | 1 (0,0) | 0,4 |  |
| Mixed | 77 (0,3) | 649 (9,2) | 2 297 (42,6) | 413 (3,1) | 16 (0,1) | 1,9 |  |
| Negative | 24 702 (97,4) | 6 383 (90,1) | 1 662 (30,8) | 143 (1,1) | 5 (0,0) | 29,1 |  |
| Very negative | 530 (2,1) | 0 (0,0) | 0 (0,0) | 0 (0,0) | 2 (0,0) | 0,5 |  |
| **Response personalisation** |  |  |  |  |  |  | <0,001 |
| High | 14 820 (58,4) | 4 223 (59,6) | 3 068 (56,9) | 4 861 (36,3) | 5 103 (33,8) | 41,8 |  |
| Medium | 1 916 (7,6) | 745 (10,5) | 519 (9,6) | 567 (4,2) | 697 (4,6) | 5,7 |  |
| Low | 8 631 (34,0) | 2 117 (29,9) | 1 801 (33,4) | 7 948 (59,4) | 9 320 (61,6) | 52,5 |  |
| **Appropriateness of response tone** |  |  |  |  |  |  | <0,001 |
| Appropriate | 15 793 (62,3) | 4 887 (69,0) | 3 962 (73,5) | 10 532 (78,7) | 12 811 (84,7) | 77,7 |  |
| Partially appropriate | 6 556 (25,8) | 1 718 (24,2) | 1 255 (23,3) | 2 761 (20,6) | 2 291 (15,2) | 18,8 |  |
| Inappropriate | 3 018 (11,9) | 480 (6,8) | 171 (3,2) | 83 (0,6) | 18 (0,1) | 3,5 |  |
| **Response category** |  |  |  |  |  |  |  |
| More frequent at high ratings |  |  |  |  |  |  |  |
| **Team appreciation** | 17 (0,1) | 16 (0,2) | 44 (0,8) | 267 (2,0) | 551 (3,6) | 2,4 | <0,001 |
| **Thank you** | 407 (1,6) | 275 (3,9) | 1 143 (21,2) | 10 406 (77,8) | 14 299 (94,6) | 64,3 | <0,001 |
| More frequent at low ratings |  |  |  |  |  |  |  |
| **Dispute / reporting** | 382 (1,5) | 42 (0,6) | 14 (0,3) | 1 (0,0) | 0 (0,0) | 0,4 | <0,001 |
| **Defence / justification** | 5 350 (21,1) | 914 (12,9) | 335 (6,2) | 85 (0,6) | 7 (0,0) | 6,1 | <0,001 |
| **Apologies** | 8 669 (34,2) | 2 874 (40,6) | 1 511 (28,0) | 562 (4,2) | 41 (0,3) | 11,5 | <0,001 |
| **Invitation to private exchange** | 994 (3,9) | 232 (3,3) | 179 (3,3) | 77 (0,6) | 3 (0,0) | 1,3 | <0,001 |
| **Factual correction** | 2 337 (9,2) | 484 (6,8) | 326 (6,1) | 206 (1,5) | 24 (0,2) | 3,0 | <0,001 |
| **Explanation of regulations** | 4 947 (19,5) | 1 385 (19,5) | 929 (17,2) | 740 (5,5) | 33 (0,2) | 6,7 | <0,001 |
| **Education on the role** | 887 (3,5) | 247 (3,5) | 168 (3,1) | 79 (0,6) | 21 (0,1) | 1,2 | <0,001 |
| **Other** | 58 (0,2) | 9 (0,1) | 5 (0,1) | 2 (0,0) | 6 (0,0) | 0,1 | <0,001 |
| **Improvement** | 632 (2,5) | 442 (6,2) | 559 (10,4) | 667 (5,0) | 34 (0,2) | 1,6 | <0,001 |
| **Brief generic response** | 598 (2,4) | 127 (1,8) | 90 (1,7) | 103 (0,8) | 83 (0,5) | 1,1 | <0,001 |
| **Practical information** | 89 (0,4) | 38 (0,5) | 85 (1,6) | 181 (1,4) | 18 (0,1) | 0,3 | <0,001 |

<sup>1</sup> unweighted n (%)
<sup>2</sup> p-value : χ<sup>2</sup> Pearson’s trend (weighted logistic regression): Rao & Scott adjustment
<sup>3</sup> Weighted total: percentages weighted by the inverse of the sampling fraction for each rating (disproportionate stratified sampling).

### Detection of AI-generated reviews

A binary classifier (human *vs* AI) was obtained by fine-tuning pre-trained BERT encoders on a balanced corpus sampling from 103 338 human reviews published before 2021 (prior to the public large adoption of ChatGPT (20)) and 80 000 reviews generated by four open-source LLMs, either created de novo (80 %) or rewritten from human reviews (20 %). The corpus was split by pharmacy into training, validation, and test sets (70/15/15 %). Six encoders were compared. To limit incorrect AI attributions, the decision threshold was calibrated to aim for a false positive rate of 1 %. The selected model, BERT large, was applied to reviews published after 2021, and the proportion of flagged reviews for each pharmacy was compared to this baseline (Supplementary Methods S4).

### Estimation of removed one-star reviews

In Germany, Google Maps indicates in brackets the number of reviews removed following complaints of defamation. We used aggregated data (rating histogram and mention of removal) from 3 430 German pharmacies to construct and validate a model of the expected distribution of ratings in the absence of removal. The expected number of one-star reviews for each pharmacy was predicted from two-to-four-star reviews (anchor category) using a Dirichlet-multinomial model fitted on listings without a mention of removal, and then compared to the observed number (deficit score). The model was validated by comparing listings with and without mentions, and then applied to French pharmacies (Supplementary Methods S5).

### Software

Data collection was carried out using Python (Play-wright), LLM coding with Ollama (Version 0.40.2) https://github.com/ollama/ollama, the classifier with the Transformers library, and statistical analyses with R (version 4.6.1). The significance threshold was set at 5 %. The code and prompts are available at https://github.com/jgodet/avisPharma.

### Ethical and regulatory aspects

The study focused exclusively on content made public by its authors and did not involve human subjects as defined by research regulations. The names of the review authors were removed from the working file. No reviews or responses are reproduced in a form that allows the identification of a user or a pharmacy.

## Results

### Data description

In total, 729 520 reviews concerning 19 985 community pharmacies were analysed. Among all the reviews, 278 504 (38.2 %) had received a response from the pharmacy team and 67.5 % of the pharmacies had responded at least once.

The number of comments per pharmacy varied strongly by region (Figure 1A). The highest medians were observed in Île-de-France (32 comments per pharmacy; IQR 0–58), Provence-Alpes-Côte d’Azur (27; IQR 7–51) and Auvergne-Rhône-Alpes (26; IQR 10–47). The lowest were observed in Pays de la Loire (12; IQR 0–35) and Bourgogne-Franche-Comté (15; IQR 5–28), and across the overseas regions (medians between 6 and 9). The distribution was highly skewed, with some pharmacies accumulating several hundred comments (up to 1 540 in Île-de-France). The distribution of ratings was bimodal, with a characteristic J-shape and a very clear imbalance in favour of positive reviews (Figure 1B). Five-star reviews represented 69 % of the reviews (506 956) and one-star reviews 18 % (130 562), while intermediate ratings were rare: two stars 3 %, three stars 2 %, four stars 8 %. The average rating was 4.08/5 and the median rating was 5/5. Although increasing, the flow of reviews remains generally quite modest, with median numbers of a few comments per year per pharmacy (Figure 1C).

**Fig. 1.**
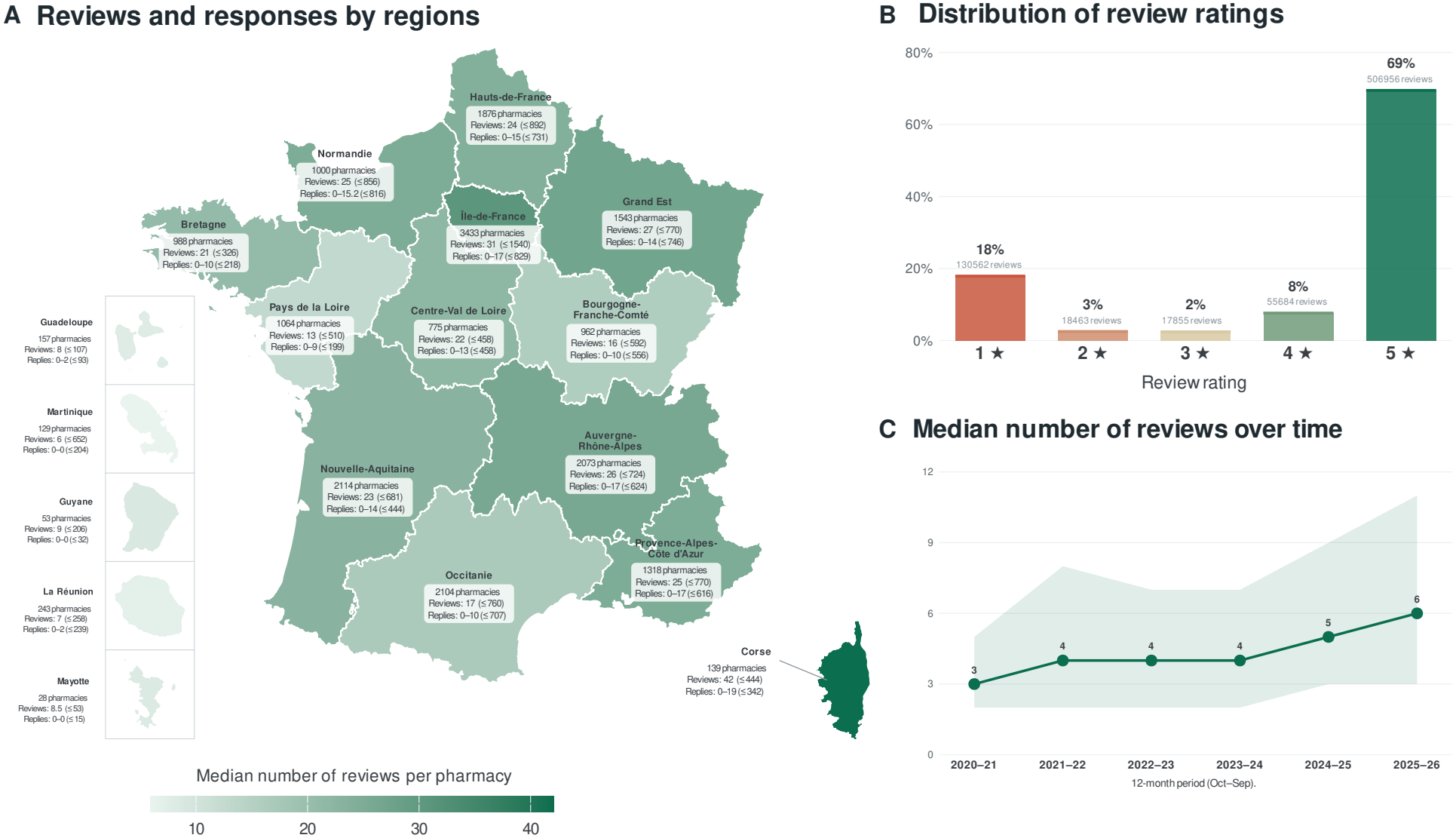
Description of Google reviews left for community pharmacies and pharmacy responses. **(A)** Median and maximum number of reviews per pharmacy, IQR of the number of responses and maximum number of responses, by region. Each region is annotated with its number of pharmacies, the median and maximum number of comments per pharmacy, as well as the interquartile range (IQR, Q1–Q3) and the maximum number of responses. Overseas regions are presented in insets, without regard to scale or geographical position. **(B)** Distribution of ratings assigned (1 to 5 stars) across all reviews (*n* = 729520; average rating 4.08/5; median 5/5). **(C)** Evolution of the median number of reviews per pharmacy and per year between 2020 and 2026 (October to September periods), Band: IQR (Q1–Q3) between pharmacies.

### Response rate and response length

Among reviews containing a comment, the response rate was 38 % for France as a whole, and the median length of responses was 21 words (Figure 2A). The metropolitan regions formed a homogeneous group, with response rates between approximately 33 and 43 % and median lengths of 18 to 25 words. The overseas regions were distinguished by lower response rates (approximately 13 % in Mayotte, 22 % in Martinique, 28 % in Guadeloupe and 31 % in La Réunion), but by longer responses (median lengths of 27 to 37 words). The width of the confidence intervals, however, calls for caution for these regions due to their small sample sizes. The raw response rate varied little according to the review rating (Figure 2B): it was 38 % for one-star reviews, 39 % for two stars, 33 % for three stars, 32 % for four stars and 39 % for five stars. Reviews with intermediate ratings therefore received a response slightly less often. This profile was generally found in most regions, with significant dispersion between regions. In contrast, the length of responses decreased markedly with the rating (Figure 2C): the median fell from 44 and 42 words for one- and two-star reviews to 34 words for three stars, and then to 18 and 17 words for four and five stars.

**Fig. 2.**
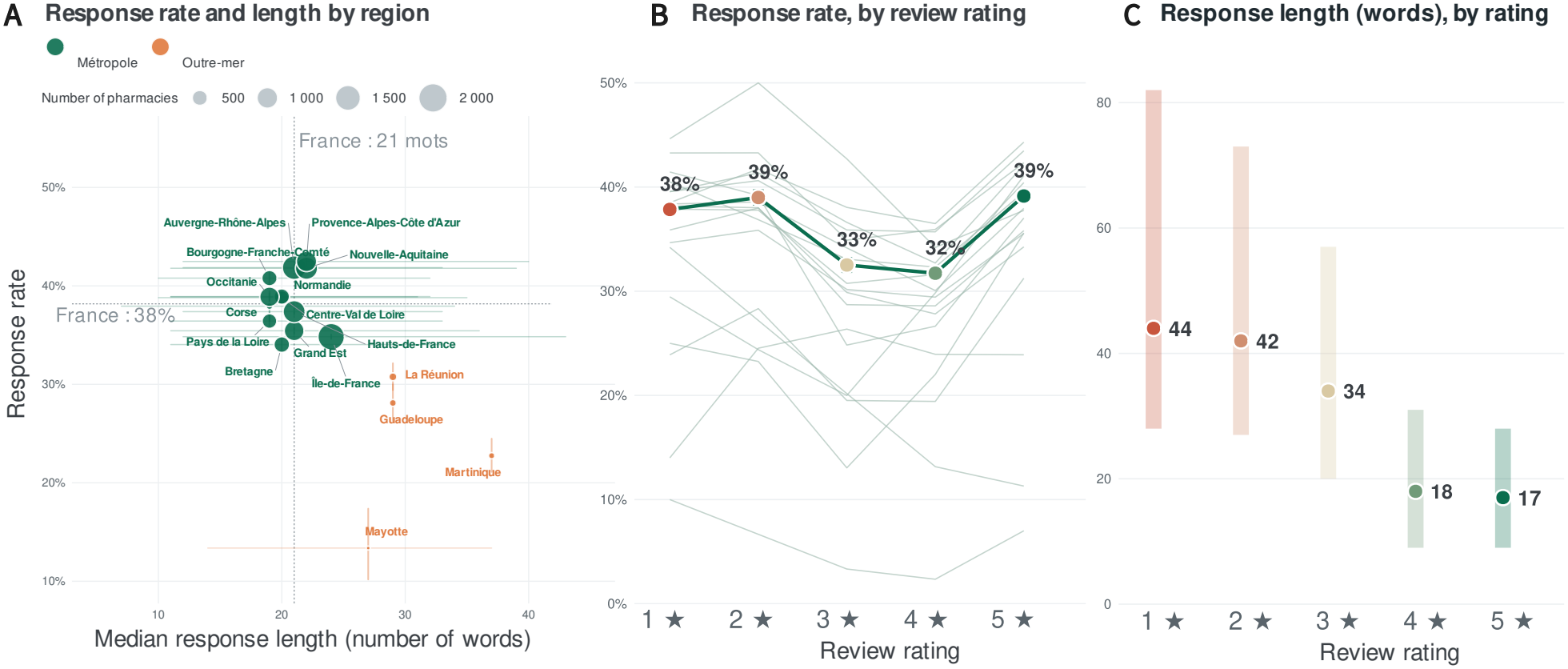
Description of pharmacy responses to reviews. **(A)** Response rate to reviews as a function of the median length of responses (number of words), by region. The bubble area is proportional to the number of pharmacies. The horizontal line represents the IQR of the response length; the vertical line, the 95 % confidence interval (95 % CI, Wilson method) of the response rate. The dotted lines indicate the values for France as a whole (response rate 38 %; median length 21 words). Green: Métropole (Mainland); orange: Outre-mer (Overseas). **(B)** Response rate according to the review rating, for France as a whole (thick line, with 95 % CI). The light lines each represent a region (regions with at least 30 reviews for the rating considered). **(C)** Length of responses (number of words) according to the review rating: median (dot) and IQR (bar). For the three panels, the denominator consists of reviews containing a textual comment. Raw, unadjusted indicators.

### Factors associated with response

For equal characteristics and within the same pharmacy, negative reviews (one or two stars) were more frequently responded to than positive reviews (five stars) (adjusted OR 1.35; 95 % CI [1.33–1.38]). The probability of response increased with the length of the review: a doubling of the number of words was associated with a 17 % increase in the odds of response (OR 1.17; 95 % CI [1.16–1.18]). Conversely, it decreased with the age of the review, with the odds of response falling by 19 % per year (OR 0.81; 95 % CI [0.81– 0.82]). Higher-rated pharmacies responded more often: each additional point of average rating multiplied the odds of response by 2.51 (95 % CI [2.27–2.77]). Regional differences persisted after adjustment. Compared to Auvergne-Rhône-Alpes (reference), the propensity to respond was lower in most regions, particularly in Corsica (OR 0.46; 95 % CI [0.25–0.86]), Grand Est (OR 0.61; [0.50–0.76]), Centre-Val de Loire (OR 0.66; [0.50–0.87]), Brittany (OR 0.69; [0.54– 0.90]), and Hauts-de-France (OR 0.71; [0.58–0.87]). The gap was more moderate in Normandy (OR 0.75; [0.58– 0.97]), Île-de-France (OR 0.79; [0.66–0.95]), Occitanie (OR 0.80; [0.65–1.00]), and Nouvelle-Aquitaine (OR 0.81; [0.66– 0.99]). Pharmacies in Provence-Alpes-Côte d’Azur, Pays de la Loire, and Bourgogne-Franche-Comté did not differ significantly from those in the reference region. The most significant effect, however, remained that of the pharmacy itself. The variance of the random intercept was very high (*σ*^2^ = 8.24), which corresponds to a median odds ratio of 15.5: for the same review, the odds of response differed by a median factor of 15.5 between two randomly selected pharmacies. The proportion of response variability attributable to the pharmacy (intraclass correlation coefficient) reached 71 %. Compared to this heterogeneity, the effects of review polarity, length, and age were modest. The decision to respond therefore seems to depend primarily on a policy specific to each pharmacy.

### Characterisation of responses

In total, a sample of 66 336 review–response pairs containing a textual comment were characterised (Supplementary Methods S3.4): 25 367 for one-star reviews, 7 085 for two stars, 5 388 for three stars, 13 376 for four stars and 15 120 for five stars (Table 1). The overall percentages presented below are weighted to account for stratified sampling and are representative of all responses to commented reviews.

The sentiment attributed to the reviews by the language model was consistent with the rating: 99.5 % of one-star reviews and 90.1 % of two-star reviews were judged as negative or very negative, and 95.5 % of four-star reviews and 99.8 % of five-star reviews were judged as positive. Three-star reviews were the most heterogeneous: 42.6 % were judged as mixed, 30.8 % negative, 15.3 % positive and 11.2 % neutral. Across all ratings, thanking was by far the most frequent primary function of the responses (64.3 %). This was followed by apologies (11.5 %), explanations of regulations (6.7 %), and defence or justification (6.1 %). Other functions each represented less than 3 % of the responses. The function of the responses varied strongly according to the rating (all trends with *p <* 0,001). Responses to positive reviews were almost exclusively limited to thanking (94.6 % of responses to five-star reviews and 77.8 % to four-star reviews), supplemented to a small extent by the valuation of the team (3.6 % for five stars). Responses to negative reviews were more diversified. For one-star reviews, the primary function was most often an apology (34.2 %), a defence or justification (21.1 %), an explanation of regulations (19.5 %), or a factual correction (9.2 %). Apologies were even more frequent for two-star reviews (40.6 %), while defence was less present there (12.9 %). The announcement of a concrete improvement remained rare, but peaked for mixed three-star reviews (10.4 %, compared to 2.5 % for one star). Challenges to the authenticity of the review (1.5 % for one star), invitations to continue the exchange privately (3.9 %), and explanations regarding the role of the pharmacist (3.5 %) were infrequent, and almost absent for positive reviews. Practical information, rare regardless of the rating (0.3 % in total), was slightly more frequent for three and four-star reviews, without a monotonic trend.

According to the automated coding, 77.7 % of the responses had a tone appropriate to the review, 18.8 % a partially appropriate tone and 3.5 % an inappropriate tone (Figure 3). The proportion of inappropriate responses increased markedly as the rating decreased: 0.1 % for five-star reviews, 0.6 % for four stars, 3.2 % for three stars, 6.8 % for two stars and 11.9 % for one star (*p <* 0.001). Thus, approximately one in eight responses to a one-star review was judged as inappropriate, and one in four only partially appropriate (25.8 %). Personalisation followed the opposite profile. Responses to negative or mixed reviews more frequently incorporated specific elements of the review (strong personalisation: 56.9 to 59.6 % for one to three-star reviews) than responses to positive reviews (33.8 to 36.3 % for four and five stars). The latter were mostly poorly personalised (59.4 to 61.6 %). In total, more than half of the responses (52.5 %) were weakly personalised.

**Fig. 3.**
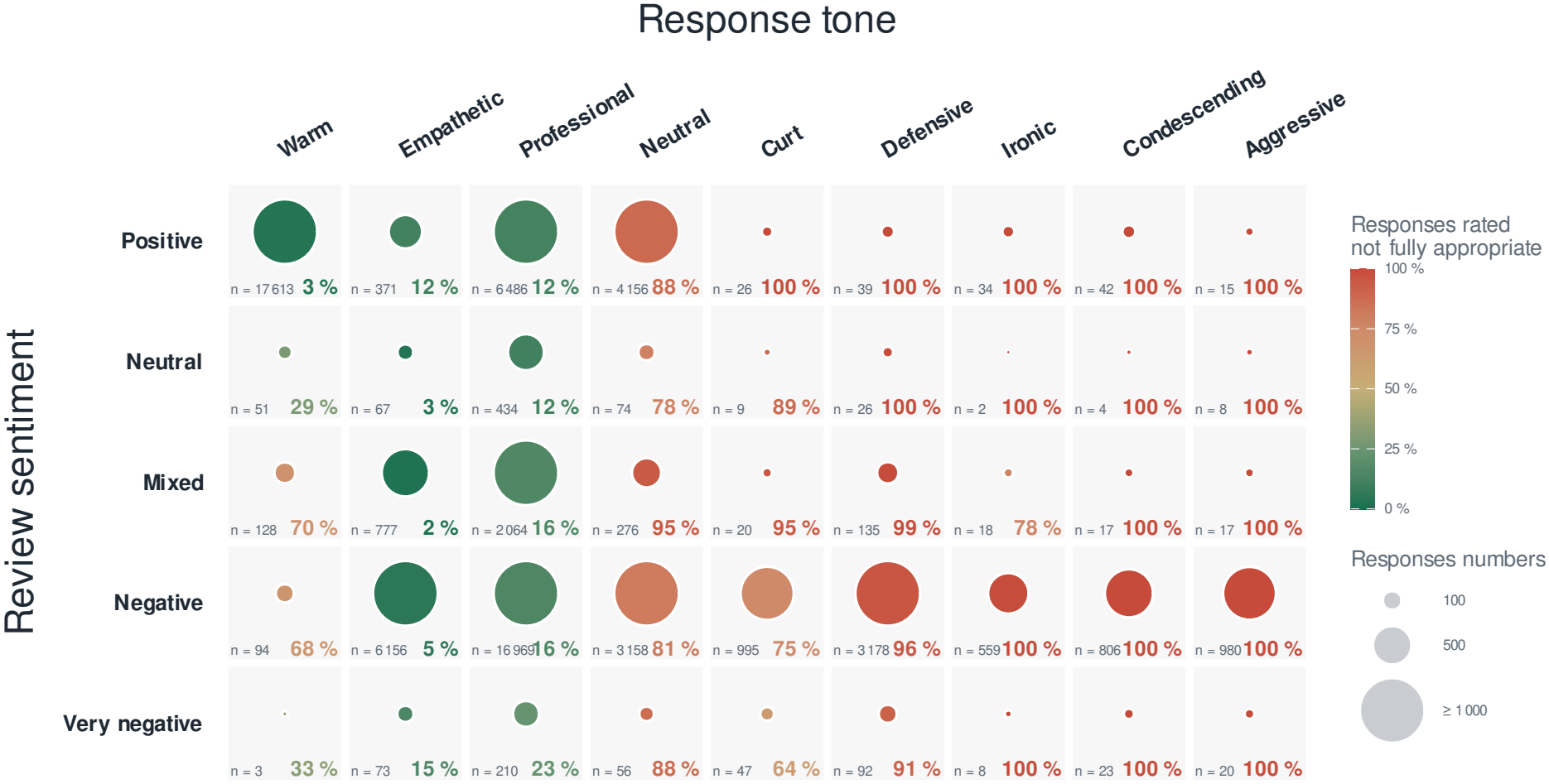
Response adequacy according to the review sentiment and the response tone. Each cell crosses the review sentiment (rows) and the tone of the pharmacist’s response (columns), as coded by the language model. The area of the bubbles is proportional to the number of review–response pairs (capped at 1 000). Their color indicates the proportion of responses judged as not adequate, i.e., inadequate or partially adequate, from green (0 %) to red (100 %). Each cell shows the count (bottom left) and the percentage of non-adequate responses (bottom right). Analysis limited to reviews containing a textual comment; unweighted counts and percentages of the sample (*n* = 66336). Empty cells: combination absent.

In summary, pharmacies responded to positive reviews with brief and standardised thanks. Their responses to negative reviews were longer, more personalised and more varied in their functions, but also more frequently defensive and of an inappropriate tone.

### Detection of reviews written by LLMs

#### AI-written review detector

A classifier was trained on two classes (human *vs* AI) (Figure 4A) on a training set of 20 000 reviews sampled from human reviews (reviews written before 2021) and reviews generated by four open models: Mistral 7B, Qwen 2.5 7B, Llama 3.1 8B, and Gemma 4 12B (Supplementary S4). The decision threshold was calibrated on the validation set for a false positive rate of 1 %. Six encoders were compared, each with five random seeds: CamemBERT (base and large), CamemBERTa-v2, ModernCamemBERT, EuroBERT-210m, and UltraBERT-150M (Supplementary Table S1 and Supplementary Figure S3). The best performing, camemBERT large, achieved an AUC of 0.99 on the test set. At the learned threshold, its sensitivity is 90.4 % with an observed false positive rate of 1.05 %. As expected, the prediction is less performant on short reviews (*≤* 15 words) and on reformulations (Supplementary Figure S2).

**Fig. 4.**
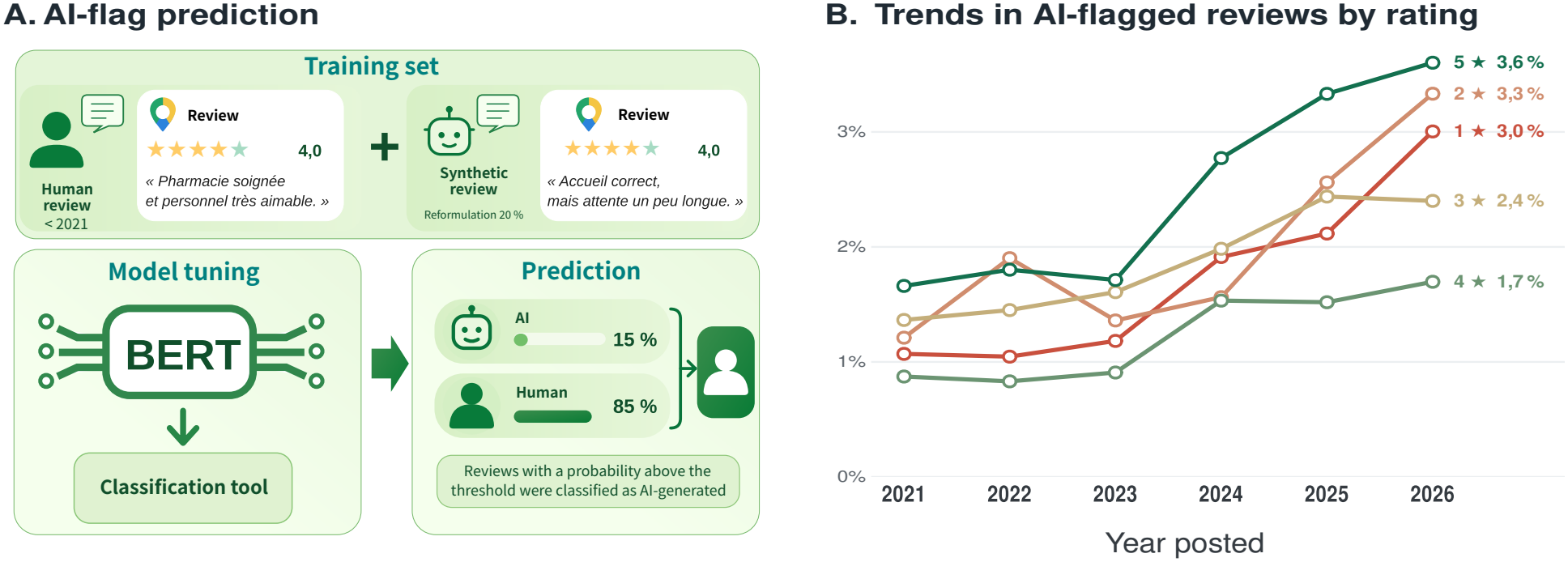
Detection of AI-generated pharmacy reviews using a fine-tuned BERT-based classifier. Proportion of pharmacy reviews flagged as AI-generated, by approximate year of posting and by star rating. Reviews were scored by a CamemBERT large classifier fine-tuned to distinguish human-written from AI-generated reviews (Supplementary Section S4). A review was flagged when its predicted probability exceeded a decision threshold set at the 99^th^ percentile of probabilities for human-written reviews in the validation set, corresponding to an expected false positive rate of about 1%. This rate is the baseline expected in the absence of any AI-generated reviews; proportions consistently above it suggest the presence of generated content. The year of posting is approximate because it was derived from the relative dates displayed by Google (e.g. “2 years ago”). Year *×* rating combinations with fewer than 50 reviews are not shown (*N* = 625,585 reviews).

#### Evolution of the proportion of flagged reviews by rating

The classifier was applied to 625 585 pharmacy reviews (Figure 4B). From 2021 to 2023, the proportion of flagged reviews remained between 0.8 and 1.9 % depending on the rating, close to the false positive rate set during calibration. It subsequently increased for all ratings starting from 2024. In 2026, it reached 3.6 % for 5-star reviews, 3.3 % for 2-star, 3.0 % for 1-star. The increase appears slightly stronger for 5-star reviews and for 1- and 2-star reviews. Intermediate ratings remained the least affected. AI is therefore likely used primarily at the two extremes of the scale, for both glowing reviews and complaints. It should be noted that these rates combine true and false positives and therefore do not directly measure actual prevalence. The latter clearly remains modest. Finally, the years of posting are approximate, as they were reconstructed from time-classes since publication displayed by Google, and the year 2026 is incomplete.

### One-star review deficit

#### Sample of German pharmacies

To study one-star review deficits—a proxy for pharmacy team intervention to manipulate ratings—we analyzed the rating histogram of German pharmacies. In Germany, Google displays on each business profile a range indicating how many reviews were removed over the past 365 days following defamation complaints (e.g., “11 to 20 reviews”). This provides an external benchmark of review removal against which we can validate a deficit score. We fitted a model to the German data and applied it to French pharmacy data to explore potential rating manipulation.

Among the data of the 3 430 German pharmacies collected, the histogram was most often reconstructed from the length of its bars (89 %), with a resolution of up to 488 reviews per bar unit; 335 of these listings (10.0 %) displayed a removal notice. The main analysis was restricted to the 1 819 pharmacies whose histogram resolution was at most 5 reviews per bar unit and which had at least one 2- to 4-star review. These pharmacies had a median of 125 reviews (interquartile range 82–178), and 232 of them (12.8 %) displayed a removal notice.

#### Reference model

We built a Dirichlet-multinomial regression model that learns from pharmacies without a notice how many one-star reviews are expected relative to the number of 2- to 4-star reviews, a category assumed to be unaffected by removals (Supplementary S5). In this model, the 5-star reviews, genuine or not, play no role. For each pharmacy of the sample, the observed number of one-star reviews is compared with this expectation, and the resulting mid-*p* value is converted into a *z*-score that follows a standard normal distribution in the absence of removals.

Fitted on the 1 587 pharmacies without a notice, the *z*-scores of reference pharmacies were centred close to zero (Figure 5A) but the model indicated strong between-pharmacy heterogeneity in the proportion of one-star reviews. Their mid-*p* values were not uniformly distributed (Figure 5B): values below 0.25 were under-represented and values between 0.55 and 0.85 over-represented. Accordingly, only 3.4 % of reference pharmacies had *p <* 0.05, fewer than the nominal 5 %. In addition, the *z*-score increased with the number of reviews (Spearman *ρ* = 0.42 in the reference population), including among pharmacies whose histogram gave one-star counts similar to the one expected (*ρ* = 0.54). From these observations, two conclusions follow. First, the test is conservative in the lower tail, so the deficits reported below are unlikely to be overstated. Second, the model overestimates the expected proportion of one-star reviews for small listings and underestimates it for large ones, so individual scores are not directly comparable between listings of very different sizes. Comparisons between populations were therefore also made within a common volume band (see below).

**Fig. 5.**
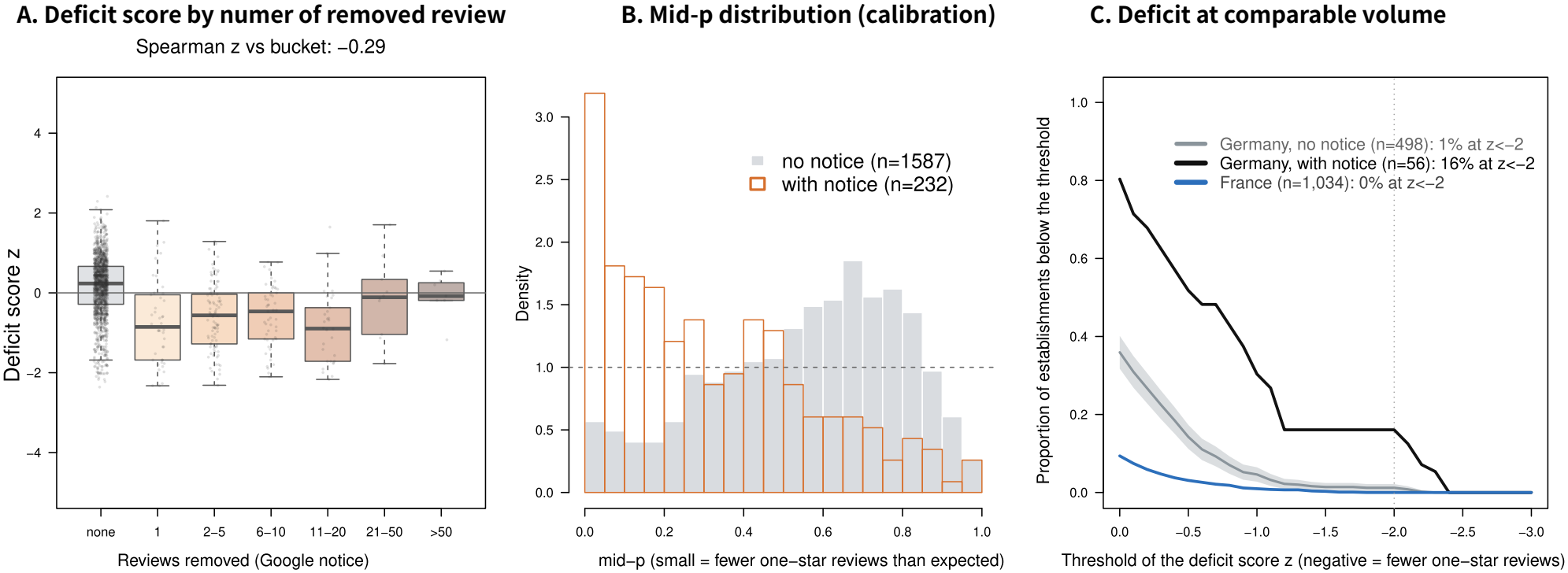
Detection of a one-star review deficit: validated in Germany, applied in France. Proportion of establishments with a one-star review deficit score *z* below each threshold (negative *z* = fewer one-star reviews than expected), compared at comparable review volume (one-star plus two-to-four-star anchor; 31–62 reviews). The grey curve, with its 95 % binomial confidence band, is the reference “no removal” population (German pharmacies with no review-removal notice); the black curve is the known-manipulated population (German pharmacies displaying a Google review-removal notice); the blue curve is French pharmacies, for which no such notice exists. The excess of the lower tail for the German with-notice population (30.6 % below *z* = *−* 2, versus 9.4 % in the no-notice reference) validates the detector at the population level; the French population shows no such excess (0.0 % below *z* = *−* 2), indicating no detectable systematic removal of one-star reviews. The vertical dashed line marks *z* = *−* 2. Counts: Germany without notice *n* = 860, with notice *n* = 111, France *n* = 1,034.

#### Deficit and Google notice

Pharmacies displaying a removal notice had fewer one-star reviews than expected: median *z*-score *−* 0.81 versus 0.1 for pharmacies without a notice, and median deficit rate 0.64 versus *−* 0.04 (one-sided Wilcoxon test, *p* = 3.2 *×* 10*−*32). Their mid-*p* values were concentrated near zero, in contrast with the reference population (Figure 5B). Pharmacies with a notice had slightly fewer reviews (median 20 versus 28 onestar or 2- to 4-star reviews), but the contrast persisted after adjustment for the number of reviews (mean difference in *z*-score *−* 0.61, standard error 0.05, *p* = 5 *×* 10*−*30) and was of similar magnitude in each volume quartile (difference in medians 0.61 to 0.71). The *z*-score was correlated with the bracket displayed by Google (Spearman *ρ* = *−* 0.29; Figure 5A), but without a gradient beyond the first removed review: median *z*-scores were similar from the 1 to the 11–20 brackets (median deficit 4.3 to 6.0 reviews), and no deficit was apparent above 20 removed reviews, brackets that included only 17 pharmacies. At the individual level, 91 pharmacies (5.0 %) had *p <* 0.05: 37 of the 232 pharmacies with a notice (15.9 %) and 54 of the 1 587 without (3.4 %). The test therefore does not allow individual establishments to be identified, but it reveals a deficit at the population level.

#### Comparison with France

To compare populations at similar review volume, German and French listings were restricted to those with 31 to 62 one-star plus 2- to 4-star reviews (Figure 5C). Within this band, at every threshold, the proportion of pharmacies below the threshold was highest among German pharmacies with a notice, intermediate among German pharmacies without a notice, and lowest among French pharmacies. Below *z* = *−* 2, these proportions were 16 % (*n* = 56), 1 % (*n* = 498) and 0 % (*n* = 1034), respectively. French pharmacies, for which Google displays no removal notice, thus showed no population-level one-star deficit: their curve lay below the confidence band of the German reference at all thresholds.The lower French curve also reflects a higher baseline share of one-star reviews in France, possibly indicating cultural differences in ratings.

#### Sensitivity analyses

The contrast between pharmacies with and without a notice was unchanged:

- when each listing was assigned the maximum number of one-star reviews compatible with the resolution of its histogram (median *z*-score *−* 0.67 versus 0.13, *p* = 5.4 *×* 10*−*30; 2.0 % of reference pharmacies with *p <* 0.05);
- when restricted to the 385 listings whose histogram gave exact counts, 59 of them with a notice (median *z*-score *−*0.71 versus 0.06, *p* = 3.5 *×* 10*−*10);
- when all 3 350 listings were included, without a resolution criterion (median *z*-score *−*0.66 versus 0.14, *p* = 6.6 *×* 10*−*48);
- when the anchor category was restricted to 3- and 4-star reviews (Spearman correlation of deficits with the main analysis *ρ* = 0.98).

Taken together, these results show no evidence of large-scale one-star review removal in French pharmacies. Removals limited to a few pharmacies or a few reviews would, however, remain undetectable at the population level.

## Discussion

### Main results

This study describes, on a national scale, the way in which French pharmacies respond to reviews published on Google Maps. Out of nearly 730 000 reviews from pharmacies across the entire country, slightly more than one in three reviews (38 %) had received a response, and two in three pharmacies had responded at least once. The decision to respond depended primarily on the pharmacy itself: the pharmacy explained 71 % of the response variability, far ahead of the characteristics of the review. For a given pharmacy, negative reviews received a response slightly more often than positive reviews, and responses approximately three times longer. Responses to positive reviews were mostly brief and little personalised thank-you messages. Responses to negative reviews employed more varied registers: apologies, justification, and explanation of regulations. According to the automated coding, the tone was appropriate in more than three-quarters of cases, but approximately one in eight responses to a one-star review was judged inappropriate. The proportion of reviews flagged as written by an artificial intelligence remained close to the detector’s background noise until 2023, then doubled in 2026, while remaining modest, in line with other areas of online health (21). Finally, the analysis of German data, where Google flags removed reviews, shows that these removals leave a measurable trace in the distribution of ratings. This trace was not detectable across the population of French pharmacies.

### Heterogeneous appropriation

The observed response rate indicates that responding to reviews has become a common practice in pharmacies, although it is not generalised: one third of pharmacies had never responded. The heterogeneity between pharmacies is the most striking result. For the same review, the response rate varied by a median factor of 15 between two randomly selected pharmacies. Responding is therefore less a reaction to the content of the review than a policy specific to each pharmacy: some respond to almost all reviews, others to none. This policy may reflect the time available, the digital appetite of the team, the size of the pharmacy, or the use of an external provider. Our data do not allow us to distinguish between these explanations.

The highest-rated pharmacies responded more frequently. Several interpretations are possible and are not mutually exclusive: an attention paid to the online image that translates into both responses and a better rating, a specific effect of responses on subsequent reviews, or common characteristics (size, location, organisation) favouring both. The crosssectional design does not allow for a definitive conclusion. Similarly, the decrease in response with the age of the review is consistent with a recent diffusion of the practice, but also with the absence of retrospective responses to old reviews.

### Responding to negative reviews

Negative reviews represent the primary challenge of responding: it is for these that a response can mitigate the effect of the review on readers (4), and it is also where the deontological risk is highest. Pharmacists seem to be aware of this: their responses to negative reviews were longer, more personalised and more frequently oriented towards explanation or apology. A portion of the dissatisfaction relates to legal constraints (prescription dispensing, substitution, reimbursement conditions) that the pharmacist cannot remove. The response then becomes a tool for education for the author and future readers. However, one in five responses to a one-star review had the primary function of defending or justifying oneself, and 12 % were judged inappropriate. A defensive or ironic response, or one that blames the customer, can damage the dignity of the profession which the code of ethics requires to be preserved (18). Above all, a response that corrects the facts carries the risk of disclosing elements of the patient’s care. The response to a review is public and permanent, and the fact that the user themselves made their visit public does not waive professional secrecy (17). Our grid did not specifically code the disclosure of information covered by secrecy (22). Factual corrections (9 % of responses to one-star reviews) and justifications nonetheless constitute the situations where this risk is most likely. Conversely, the invitation to continue the exchange privately, which allows the complaint to be handled without public exposure, remained rare (4 %).

### Influence of the emergence of conversational AI tools

The proportion of reviews flagged by the detector increased from 2024 across all ratings, reaching approximately twice the 2021–2023 level by 2026. As the threshold was calibrated for 1 % false positives, the excess attributable to reviews written or rewritten by an AI is in the range of 1 to 3 points depending on the rating. This emerging phenomenon therefore remains minority. The increase affected both ends of the scale: glowing reviews, which could correspond to complimentary reviews or satisfied users assisted in drafting, and one or two-star reviews, which suggest complaints formatted by an AI. These two uses do not have the same impact: a real review reformulated by an AI remains the testimony of an experience, whereas a review generated entirely from scratch reflects none. The detector does not allow them to be distinguished.

The same tools are likely used by pharmacies to draft responses. We have not measured this. The high proportion of poorly personalised responses could reflect either preprepared response templates or automatically generated responses. An AI-generated response raises the same ethical questions as a response written by the pharmacist, for which the pharmacist remains responsible. Even when well-configured, AI tools are not immune to deviating from the rules (23). The risk of a standardised response, or a response that indiscriminately incorporates elements of the review, should be taken into account in any future recommendations.

### Strengths and limitations of the study

This study covers almost all French pharmacies, identified using a national database, and all accessible reviews. To our knowledge, this is the first national description of pharmacists’ responses to online reviews. Mixed models allowed for the separation of effects specific to reviews from those specific to pharmacies. The coding of responses and the detection of AI-written reviews were performed using locally executed models, without data transmission to a third party. The code and prompts have been made available.

This study has several limitations. Only reviews published and visible at the time of collection were observed. Reviews filtered or removed by the platform escaped analysis, which prompted the complementary analysis of German data. Furthermore, the tone, function, and adequacy of the responses were coded by a language model and not by human evaluators. The judgment of adequacy reflects the criteria defined in the prompts, rather than the perception of users or the regulatory body. Inadequate responses may be underestimated, as the model may have judged responses that were polite but problematic regarding professional secrecy as acceptable. The detector for AI-written reviews was trained on texts produced by four open models. Its sensitivity to texts produced by commercial assistants, which are likely the most used, is unknown, and it is less performant on short reviews and reformulations. The estimation of the one-star review deficit relies on histograms largely reconstructed from bar lengths, on an assumption of stability for 2-to-4-star ratings, and on a model whose fit varies with the volume of reviews. Finally, we do not know who writes the responses (pharmacist owner, team member, service provider), nor whether responses modify subsequent reviews or the choice of users.

### Perspectives

These results argue for professional support. A professional recommendation could specify what a response may and may not contain (22). It could also help in adopting a tone appropriate for negative reviews and provide a framework for the use of AI tools to draft responses. Several complementary studies would be useful:

- a dedicated coding of potential breaches of professional secrecy or reglementation in the responses;
- a study of the perception of responses by users;
- a longitudinal follow-up on the evolution of practices, the proportion of reviews written by AI and the withdrawal of reviews;
- a qualitative survey of pharmacists regarding their motivations, their organisation and their use of service providers or AI.

## Conclusion

French community pharmacies have widely adopted the practice of responding to online reviews, but they have done so in a very heterogeneous manner: the decision to respond depends primarily on a policy specific to each pharmacy. Responses are most often tailored, remaining brief for positive reviews and more elaborate for negative reviews, yet a significant proportion of responses to the most critical reviews remain defensive or inappropriate, potentially posing a risk of breaching professional ethics. The emergence of AI-generated reviews and the possibility of having negative reviews removed further complete this picture. These results could serve to develop professional recommendations to secure these communication practices between community pharmacies and their patients.

## Data Availability

All analysis scripts are available upon reasonable request to the authors. Data cannot be shared.

## ACKNOWLEDGEMENTS

The authors thank Nedah Asas and Rym Mousli for initiating the data collection during their research visit to the laboratory.

## Funding

This study received no specific funding. The work originated as an internship project in iCube laboratory, focusing on web scraping and natural language processing for students in Master’s program in Health Data Science (Master Ingénierie de la Santé - parcours Intelligence des données en Santé) at the University of Strasbourg. DS, LM, and FH are enrolled in this program.

## Author contributions

Conception: JG; data collection: DS, LM, FH, JG; analysis: JG; drafting: JG; critical revision: DS, LM, FH, JG

## Declaration of competing interests

The authors declare no competing interests.

## Declaration of the use of generative AI and AI-assisted technologies in the writing process

During the preparation of this work, the authors utilised Claude Opus 5.5 (Anthropic) for the review of computer code, to improve the style of the manuscript, and Gemma4 model (Google) for its translation from French into English. The authors assume full responsibility for the published article.

## Supplementary Information

### S1. Source of pharmacies and collection of reviews

#### Source of pharmacies

We used the public extraction from the National File of Sanitary and Social Establishments (Fichier National des Établissements Sanitaires et Sociaux or FINESS), disseminated on the French public data open platform (data.gouv.fr) by the Ministry of Health (https://www.data.gouv.fr/api/1/datasets/r/2bb8cbe9-0291-4705-9cbc-092e6513dcc3, accessed September 9, 2026). From this listing, we extracted a list of 20 003 community pharmacies. Each pharmacy was described by an identifier (FINESS number), a name, an address, a municipality, and a region. https://www.data.gouv.fr/api/1/datasets/r/98f3161f-79ff-4f16-8f6a-6d571a80fea2

#### Automated collection

Reviews were collected between September and October 2026 using a script developed in Python (Playwright library, headless Chromium browser). For each pharmacy, a query combining the name, address elements including the municipality, and the term “pharmacy” was submitted to Google Maps. The average rating of the listing was recorded, and then the list of reviews was scrolled through until no new reviews were loaded, up to a limit of 80 scrolls. Truncated reviews were expanded before extraction.

#### Variables collected

For each review, the rating (one to five stars), the text, and the age displayed by the platform were collected, as well as the text and age of the pharmacy team’s response when available. Reviews containing a rating but no comment were also collected. The age (in months) was deduced from the relative date displayed by Google.

#### Deduplication

Duplicates were removed based on the pharmacy, the date, the note, and the text.

### S2. Analysis of responses to reviews

#### S2.1. Descriptive indicators

The descriptive analysis focused on reviews containing a textual comment. Reviews without text (rating only) or those whose text was not usable were excluded. A review was considered to have received a response when the “pharmacy team response” field contained at least one word. The length of each response was measured by the number of words, after space normalization, counting each sequence of non-whitespace characters as a word.

Indicators were calculated for all of France and by administrative region. For each region, we recorded the number of pharmacies and the number of commented reviews. The response rate was defined as the number of reviews that received a response relative to the number of commented reviews in the region; it is presented with its 95 % confidence interval (95 % CI) calculated using the Wilson method. Among reviews that received a response, the length of the responses was described by the median and the interquartile range (IQR). These indicators are descriptive and do not account for the clustering of reviews by pharmacy.

#### S2.2. Main outcome models

##### Probability of response

The unit of analysis was the review. The outcome measure was the presence of a response from the pharmacy team (yes/no). Reviews were classified as negative (one or two stars) or positive (five stars) (review polarity). The association between review polarity and the probability of response was estimated using a mixed-effects logistic regression, with a random intercept per pharmacy (FINESS number) to account for the non-independence of reviews from the same pharmacy. The model was adjusted for:

- the length of the review (logarithm of the number of words plus one);
- its age (in months, derived from the relative date displayed by Google);
- the average rating of the pharmacy;
- its region.

The model was estimated by maximum likelihood with the simplified Laplace approximation (*n*_*AGQ*_ = 0) from the lme4 package, chosen for computational time reasons given the sample size. The results are expressed as adjusted odds ratios (OR) with their 95 % Wald confidence intervals.

##### Predicted rates and risk difference

To facilitate interpretation, predicted response rates were calculated for each polarity by setting the random effect to zero (“typical” pharmacy) and averaging predictions over the observed characteristics of the reviews. We derived an adjusted risk difference (in percentage points) with a 95 % CI obtained by the delta method (marginaleffects package).

##### Response length

Response length (number of words) was compared between negative and positive reviews, among reviews that received a response, using a linear mixed model on the logarithm of the number of words, with a random intercept per pharmacy and the same adjustments. The effect is expressed as an adjusted geometric mean ratio (aGMR).

##### Software

These analyses were performed with R (version 4.6.1, 2026), using the lme4 and marginaleffects packages. The significance threshold was set at 5 %.

### S3. Automated coding of the nature and tone of the responses

#### S3.1. Categorization grid for functions

The functions of the responses were classified using a grid of thirteen categories that were mutually exclusive at the level of the main function (Table 1). This grid was constructed inductively, based on the reading of a sample of responses, and supplemented by themes described in the literature and by informal inquiries from health professionals. It covers the following registers:

- relational: thank you, valorization of the team, apologies, invitation for a private exchange;
- corrective: announcement of an improvement, factual rectification;
- informative: explanation of regulations, practical information, education on the role of the pharmacist;
- defensive: contesting the authenticity of the review, justification.

A “generic brief response” category grouped short and standardized responses without content specific to the review, and an “other” category allowed for the classification of responses that did not fit into any of the previous categories.

#### S3.2. Language model and parameterization

The coding of review sentiment and response categories was performed automatically by a large language model (LLM), and not by human coders. We used the Gemma 4 model (12 billion parameters, quantized version gemma4:12b-it-qat), run locally with the Ollama software (version 0.40.2): no data was transmitted to a third-party service. For each evaluation, the model received the review rating, its text, and the pharmacist’s response, accompanied by an instruction that defined each category and the coding rules described below. Its output was constrained to a structured format (JSON) where the possible values were limited to the planned modalities. The temperature was set to 0 to make the coding deterministic. The pairs for which the model had not returned a valid output after three attempts were resubmitted during a second pass. Reviews without a response were classified as “no response” and were not submitted to the model.

#### S3.3. Coded dimensions and coding rules

For each evaluated review–response pair, the model coded seven dimensions: two concerning the review and five concerning the response (Table S1).

##### Main and secondary functions

Each response was assigned a single main function, defined as its dominant function, and zero, one, or several secondary functions among the remaining twelve categories. The same category could not be both main and secondary: any duplicates returned by the model were removed. Thanking was to be retained as the main function only when the response was essentially limited to thanking; as soon as it also contained apologies, an explanation, or a rectification, that other function became the main one.

**Supplementary Table S1.** Dimensions coded by the language model for each review–response pair.

| Object | Dimension | Modalities |
|---|---|---|
| Review | Sentiment | positive, neutral, mixed, negative, very negative |
| Review | Tone | praising, cordial, factual, disappointed, irritated, aggressive, insulting, no text (review limited to a rating) |
| Response | Main function | one category among thirteen (table 1) |
| Response | Secondary functions | zero, one or more among the remaining twelve categories |
| Response | Tone | warm, professional, neutral, empathetic, curt, defensive, condescending, ironic, aggressive |
| Response | Appropriateness of tone to the tone and sentiment of the review | appropriate, partially appropriate, inappropriate |
| Response | Personalization | high, medium, low |

##### Tone appropriateness

A response was to be judged:

- *appropriate* when it was proportionate to the review: warm thanks for a positive review; listening, apologies, or a calm explanation for a criticism; courteous firmness in the face of an aggressive or presumed fake review;
- *partially appropriate* when it remained correct but generic, cold in the face of real disappointment, or only addressed part of the problem;
- *inappropriate* when it was defensive, ironic, condescending, or aggressive, questioned the customer, missed the point, or was a clearly off-topic copy-paste.

##### Personalization

Personalization was to be judged strong when the response incorporated specific elements of the review, and weak when it could have responded to any review.

The full prompt is provided on github.com/jgodet/avisPharma.

#### S3.4. Sampling for tone analysis

The characterization of the tone of the responses was based on a disproportionate stratified sample of 81 953 review–response pairs that received a response from the owner, with the review rating (one to five stars) defining the strata. For each rating, all responses were retained when their number was below a ceiling of 20 000. The two, three, and four-star strata were thus included in full (7 200, 5 806, and 17 667 responses, respectively), and the one- and five-star strata were undersampled (25 640 responses each, out of approximately 49 600 and 197 700 available responses). This plan aimed to ensure sufficient sample sizes for negative and mixed reviews, which are minorities in the population (2 to 3 % of reviews for two- and three-star ratings), while limiting the number of calls to the language model for the most populous strata. With these sample sizes, the half-width of the 95 % confidence interval of a proportion did not exceed 1.3 points in each stratum.

The results are presented by rating; the estimates for all responses were weighted by the inverse of the sampling fraction of each stratum:

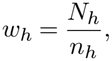

where *N*_*h*_ is the number of available responses and *n*_*h*_ is the number of sampled responses for rating *h*.

### S4. Detection of AI-generated content

To estimate the proportion of reviews written or rewritten by an LLM, we trained a binary classifier (human *vs* AI) on a corpus combining authentic reviews and generated reviews. These generated reviews included both entirely created reviews and AI-rewritten human reviews.

#### S4.1. Reference human reviews

Human reviews were drawn from the corpus by keeping only those published before 2021, more than a year before the date ChatGPT was made publicly available. Before this date, the use of an LLM to write a review can be considered negligible. The texts were normalized (Unicode NFC form, spaces and line breaks reduced to a single space). Reviews of fewer than three words and duplicates were excluded. We thus obtained 103 338 human reviews from 13 811 pharmacies.

#### S4.2. Generation of AI reviews

In total, 80 000 reviews were generated using four open-source LLMs from different families, executed locally with Ollama (version 0.40.2): Mistral 7B, Qwen2.5 7B, Llama 3.1 8B, and Gemma 4 12B (QAT version). The use of multiple model families aimed to prevent the classifier from learning the style of a single generator. For each review, the generator and a reference human review were drawn at random, and the sampling temperature was drawn uniformly between 0.6 and 1.2. Two generation modes were used:

- **Creation** (80 %) : the model wrote a Google review for a pharmacy, using the star rating of the reference human review. In 25 % of cases, the instruction was minimal, imitating a rushed user (« Write a Google review for a pharmacy, *k* stars »). In other cases, it specified a target length and one or two styles drawn from twelve: colloquial language, enthusiastic or annoyed tone, natural spelling mistakes, lack of capitalization, emojis, SMS abbreviations, elderly person unfamiliar with the internet, regular customer, mention of a specific detail (price, medication, employee’s first name), comparison with another pharmacy, etc. Target lengths (one sentence, 2–3 sentences, 4–6 sentences, a paragraph of 8–12 sentences) were drawn with probabilities of 0.51, 0.32, 0.14, and 0.03, respectively, chosen to reproduce the length distribution of human reviews.
- **Reformulation** (20 %) : the model rewrote the reference human review according to an instruction drawn from four (clarify, correct mistakes, make more professional, make more convincing). This mode simulates “AI-assisted” reviews.

##### Post-processing

The prompt requested that only the review text be produced. Typical LLM artifacts were then removed from the outputs, as they would have provided the classifier with trivial clues: introductory formulas (« Here is… »), reasoning tags, Markdown formatting, stars and ratings such as « 4/5 », and enclosing quotation marks. The same normalization as for human reviews was then applied.

#### S4.3. Constitution of datasets

From these two corpora (human and LLM), the two classes were balanced by random undersampling of the majority class, and then the corpus was split by groups into training, validation, and test sets (70/15/15 %). All reviews from the same pharmacy, as well as reformulations of a human review and their source review, always fell into the same set to avoid any information leakage between sets.

#### S4.4. Training and calibration

The classifier was obtained by fine-tuning a pre-trained encoder model for sequence classification (Transformers library, version 5.20.2). The hyperparameters are summarized in Table S2. The best checkpoint was selected based on the area under the ROC curve (AUC) in validation. Since falsely accusing a human author is the most costly error, the decision threshold was not set at 0.5: it was calibrated on the validation set at the 99^th^ percentile of probabilities assigned to human reviews, targeting a false positive rate of 1 %.

**Supplementary Table S2.** Fine-tuning hyperparameters for the human *vs* AI classifier.

| Parameter | Value |
|---|---|
| Maximum text length | 256 subword tokens (truncation) |
| Number of epochs | 3 |
| Optimiser | AdamW |
| Learning rate | 2 × 10<sup>−5</sup> (adjusted per model: 10<sup>−5</sup> to 3 × 10<sup>−5</sup>) |
| Weight decay | 0.01 |
| Warm-up | 10% of steps |
| Batch size (effective) | 16 |
| Precision | mixed |
| Random seeds | 2 per model |
| Checkpoint selection | Maximum validation AUC |
| Decision threshold | 99<sup>th</sup> percentile of probabilities for human-written reviews (validation) |

#### S4.5. Model comparison

Six encoders were compared: CamemBERT-base and CamemBERT-large (24), CamemBERTav2-base (25), ModernCamemBERT-base (26), EuroBERT-210m (27) and UltraBERT-150M (28). The learning rate was adjusted for each model (10*−*5 to 3 *×* 10*−*5; effective batch size of 16). Each model was trained with two random seeds. Models were evaluated on the test set using the following criteria:

- AUC;
- sensitivity (true positive rate) and false positive rate at the calibrated threshold;
- F1 score;
- maximum achievable sensitivity at a 1% false positive rate (optimal *a posteriori* threshold), used to assess threshold calibration;
- inference throughput.

Performance was also broken down by length band (*≤*15, 16–40, 41–100, *>*100 words), by generator and by generation mode. The selected model, CamemBERT large, achieved the highest mean sensitivity at the calibrated threshold.

#### S4.6. Application to the corpus

The selected model was then applied to reviews published after 2021, using the same normalisation. Each review was assigned a probability of being AI-generated and was flagged if this probability exceeded the calibrated threshold. Reviews shorter than 15 words, for which detection is less reliable, were identified separately. Results were aggregated at the pharmacy level: the proportion of flagged reviews for each pharmacy was compared with the expected false positive rate (approximately 1%), which represents the baseline level in the absence of any generated reviews.

### S5. Modeling of removed low-rated reviews

#### S5.1. General principle

Platforms do not publish the reviews they remove. To estimate the scale of these removals, we used a situation where some of this information is visible: in Germany, Google Maps displays a mention on an establishment’s profile indicating, in brackets, the number of reviews removed following complaints of defamation (“… *Bewertungen wurden aufgrund von Beschwerden wegen Diffamierung entfernt*”), over the last 365 days. These German data allowed for the construction and validation of a model of the expected distribution of ratings in the absence of removal. This model was then applied to French pharmacies, for which no such mention exists.

#### S5.2. Sample of German pharmacies

German pharmacies were identified using the official directory aponet.de, via a postal code search for the city centers of the 215 most populous cities in Germany. The 25 pharmacies closest to each search point were selected, and then duplicates and pharmacies without data were eliminated, totaling 3 430 pharmacies. For each, the corresponding Google Maps profile was searched using its address. When multiple profiles were suggested, the one whose name most closely resembled the aponet name was selected (string similarity after removing generic terms). Similarity matches below 0.6 were marked as doubtful and excluded.

The collection, carried out on October 31, 2026, with an automated browser (Playwright, German interface), only concerned aggregated data displayed publicly, without any review text: average rating, total number of reviews, distribution of 1 to 5-star reviews, and the text of the removal mention. This mention was converted into the bracket displayed by Google (none, 1, 2–5, 6–10, 11–20, 21–50, *>*50 reviews removed). Only profiles whose histogram contained at least 20 reviews were analyzed.

#### S5.3. Reconstitution of rating distributions

For each pharmacy, the distribution of ratings (1 to 5 stars) was recorded from the Google Maps listing, as well as any mention indicating that reviews had been removed over the last 365 days, along with the range of the number of reviews concerned (1, 2– 5, 6–10, 11–20, 21–50, *>*50). When the displayed histogram did not directly provide the counts, the bar lengths were converted into proportions and then scaled to the total number of reviews, while preserving this total (largest remainder method). The resolution of this reconstitution was measured by the number of reviews represented by one bar unit. Listings without a legible histogram were excluded. The main analysis was restricted to listings where the resolution was at most 5 reviews per bar unit and which had at least one review of 2 to 4 stars.

#### S5.4. Deficit of one-star reviews

##### Principle

Ratings were grouped into three categories: one-star reviews, an anchoring category grouping ratings from 2 to 4 stars, assumed to be minimally affected by potential deletions, and five-star reviews. The principle is to estimate, based on the anchoring category, the number of one-star reviews expected in the absence of deletions, and then to compare it to the observed number.

##### Model

The counts **n**_*i*_ = (*n*_*i*1_, *n*_*i*,anchor_, *n*_*i*5_) for pharmacy *i* were modelled using a Dirichlet-multinomial distribution:

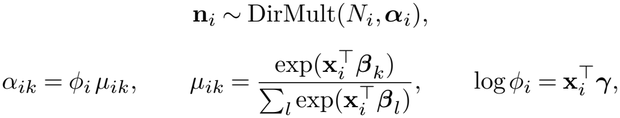

where the anchor category serves as the reference (***β***anchor = 0) and the dispersion parameter *ϕ*_*i*_ captures between-pharmacy variability beyond multinomial variability. The covariates **x**_*i*_ described the internal shape of the anchor category (smoothed log-ratios of 3-star to 2-star counts and of 4-star to 3-star counts) and its size (log(1 + *n*_*i*,anchor_)). The model was fitted by penalised maximum likelihood (*L*_2_ penalty, L-BFGS-B algorithm) on a reference population: pharmacies whose listing carried no notice of review removal. Each reference pharmacy was evaluated using leave-one-out cross-validation, i.e. with a model refitted without it.

##### Expected count and deficit score

By the aggregation property of the Dirichlet-multinomial distribution, the number of one-star reviews, conditional on the total

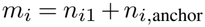

follows a beta-binomial distribution:

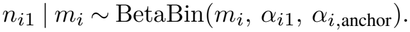

This formulation makes the estimate insensitive to any fake five-star reviews. The expected number of one-star reviews is

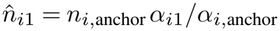

The deficit was defined as 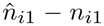 (positive when one-star reviews are missing) and the deficit rate as 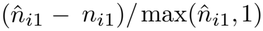. To compare listings of different sizes, a one-sided mid-*p* value,

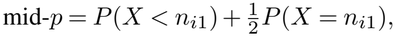

was computed under this distribution and converted into a score *z*_*i*_ = Φ*−*1(mid-*p*); a negative *z*_*i*_ indicates a deficit. The one-sided *p*-value *P* (*X ≤ n*_*i*1_) was used for individual flagging (*p <* 0.05), with Benjamini–Hochberg correction for multiple testing.

#### S5.5. Validation

Calibration was verified on the reference population: proportion of reported pharmacies, uniformity of the mid-*p* distribution, and absence of trend in the *z* score according to the number of reviews. Pharmacies with and without a Google mention were compared using a one-sided Wilcoxon test on the deficit rate and on the *z* score. The link with the range displayed by Google was measured by Spearman’s correlation. The contrast was adjusted for volume using a linear regression of the *z* score on the presence of a mention and on log(1 + *m*_*i*_), and examined by quartile of *m*_*i*_.

#### S5.6. Sensitivity analyses

Four sensitivity analyses were conducted:

1. a conservative analysis, where each page receives the maximum number of one-star reviews compatible with the resolution of its histogram (read count increased by half a bar unit);
2. a restriction to pages whose histogram gave the exact counts;
3. the analysis of all pages, without resolution criteria, using a model without covariates;
4. an anchoring category restricted to 3 and 4-star ratings, to account for potential deletions of 2-star reviews.

#### S5.7. Effect on the displayed rating

For pharmacies with a mention, a corrected rating was calculated by reintegrating the removed reviews as one-star reviews, using first the lower bound and then the upper bound of the displayed range. Since the mention covers the last 365 days and the histogram covers the entire lifespan of the listing, this correction is indicative.

##### Software

These analyses were performed with R (version 4.6.1).

---

**Supplementary Fig. S1.**
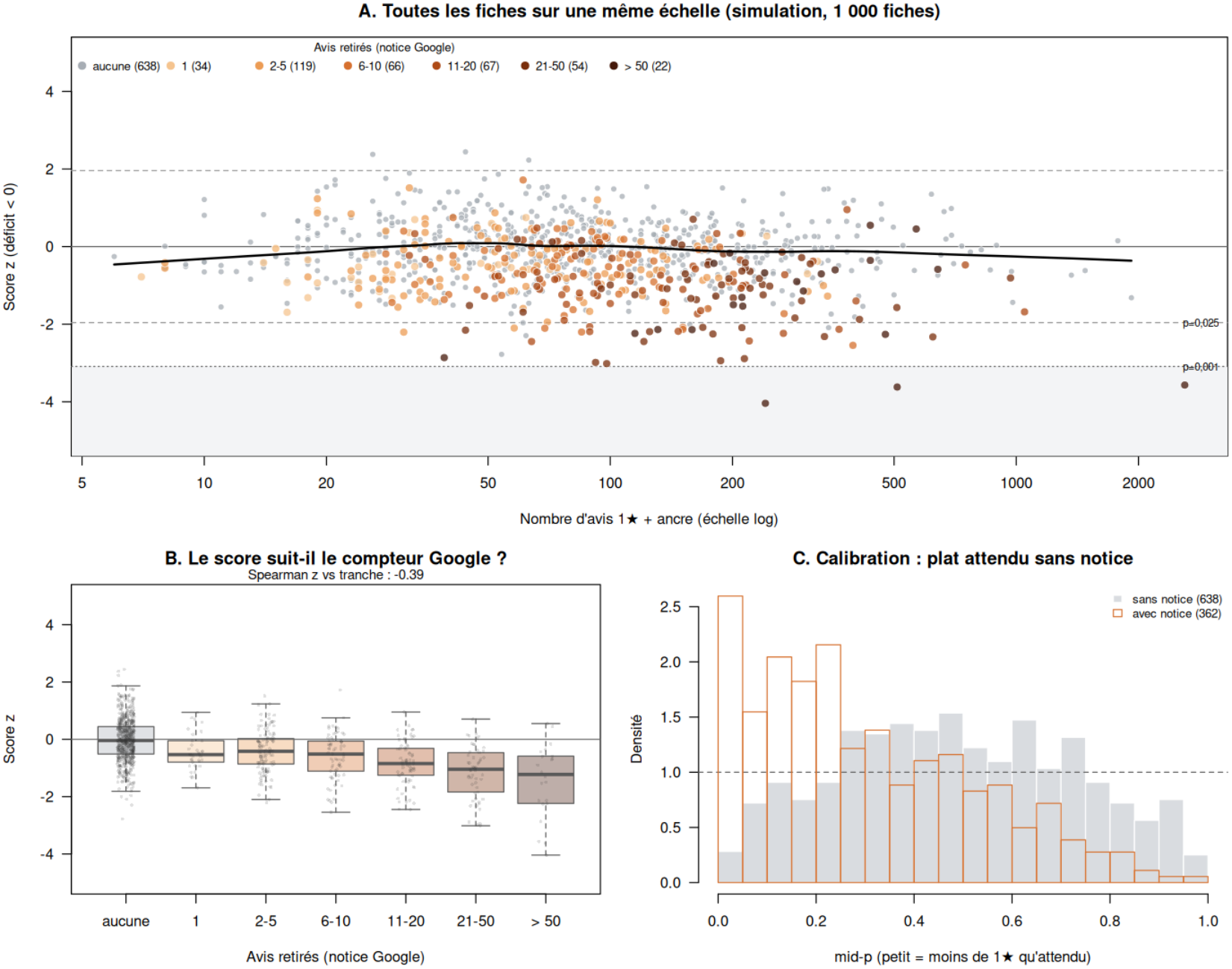
Validation of the deficit score on simulated data with known removals. Two populations of 1,000 listings were simulated using a J-shaped rating model: a mixture of a satisfied-customer component, whose propensity *a* varies across listings, and a dissatisfied-customer component whose shape is common to all listings. About 30% of listings additionally received fake five-star reviews. The total number of reviews followed a log-normal distribution (minimum of 20). The regression model does not know this generating shape, so the simulation tests its robustness. The reference population (no removals) was used to fit the model, which was then applied to the test population. In the test population, 40% of listings lost a random fraction *d* ~ Beta(2, 3) of their one-star reviews and a fraction 0.3 *d* of their two-star reviews. Only the test population is shown. **(A)** Deficit score *z* for each simulated listing as a function of the number of one-star reviews and of 2- to 4-star reviews (logarithmic scale). A negative score indicates fewer one-star reviews than expected. Colour indicates the band of the actual number of removed reviews (one- and two-star), coded in the same way as the Google notice, from grey (no removal) to dark orange; the number of listings in each band is shown in the legend. Horizontal dotted lines correspond to *p* = 0.025 and *p* = 0.975, and the shaded area to *p <* 0.001. The black curve is a locally weighted regression (lowess) of the *z* score for listings without removals; it remains flat at zero if the model is well calibrated across all volumes. **(B)** Distribution of the *z* score by band of removed reviews: median and quartiles (boxes), range excluding outliers (whiskers) and individual values (points). Scores are truncated at *±*5 for display. The Spearman correlation coefficient between *z* and the band is shown above the panel. **(C)** Distribution of mid-*p* values for listings without removals (grey) and with removals (orange outline). In the absence of a deficit, the expected distribution is uniform (dotted line, density of 1); an excess of values close to 0 reflects detection of the introduced deficit. Exact simulated counts (no histogram rounding); full model with covariates (internal shape of the anchor category and log(1 + *n*_anchor_)), without cross-validation.

**Supplementary Fig. S2.**
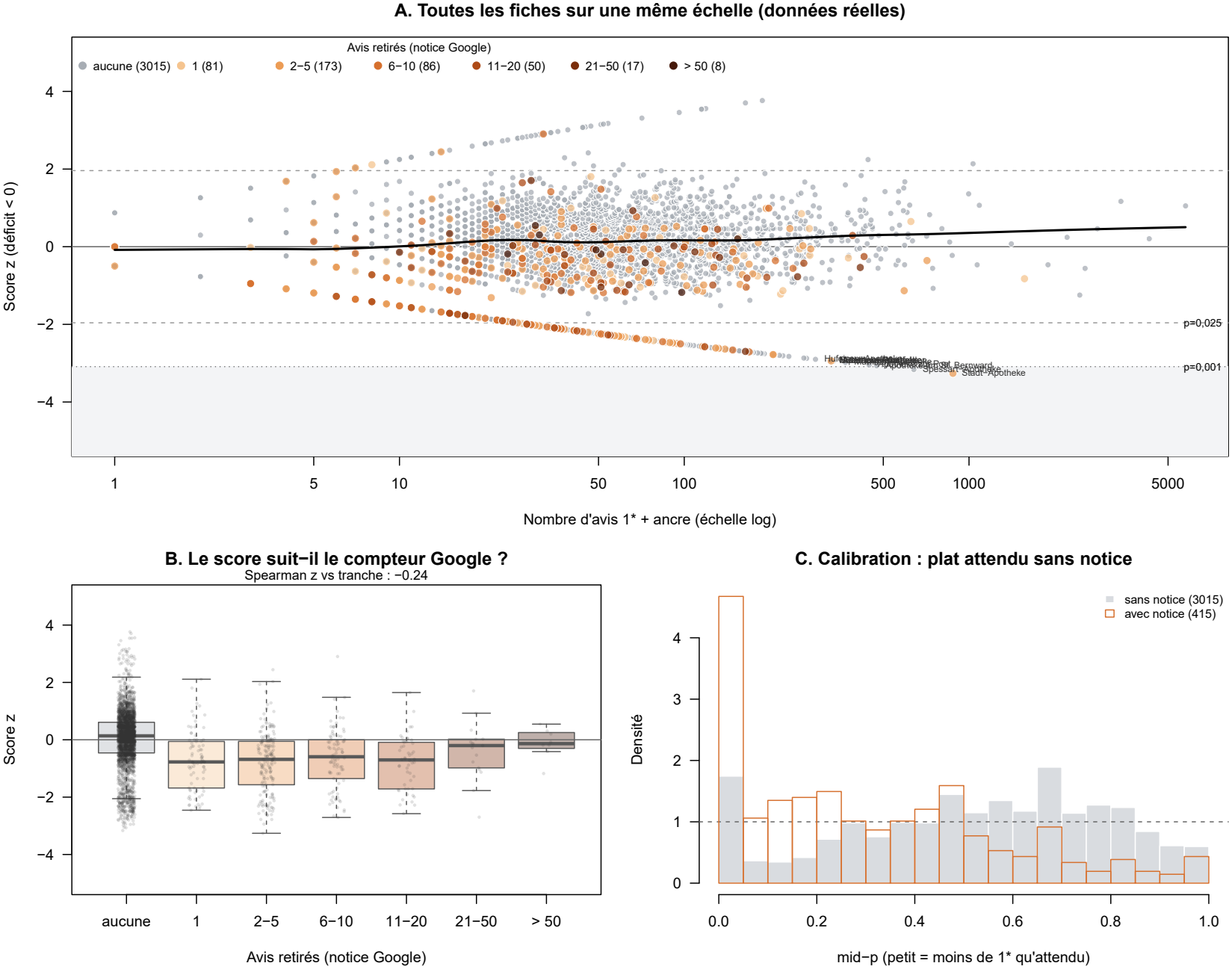
One-star review deficit by the review removal notice displayed by Google (German pharmacies). **(A)** Deficit score *z* for each pharmacy as a function of the number of one-star reviews and of 2- to 4-star reviews (logarithmic scale). A negative score indicates fewer one-star reviews than expected. Colour indicates the band of removed reviews displayed by Google, from grey (no notice) to dark orange; the number of pharmacies in each band is shown in the legend. Horizontal dotted lines correspond to *p* = 0.025 (two-sided) and the shaded area to *p <* 0.001. The black curve is a locally weighted regression (lowess) of the *z* score for pharmacies without a notice; it would remain flat at zero if the model were perfectly calibrated across all volumes. **(B)** Distribution of the *z* score by band of removed reviews: median and quartiles (boxes), range excluding outliers (whiskers) and individual values (points). Scores are truncated at *±*5 for display. **(C)** Distribution of mid-*p* values for pharmacies without a notice (grey) and with a notice (orange outline). In the absence of a deficit, the expected distribution is uniform (dotted line, density of 1); an excess of values close to 0 reflects a deficit of one-star reviews. The main analysis was restricted to the 1,819 pharmacies whose rating histogram had a resolution of at most 5 reviews per bar unit and included at least one 2- to 4-star review (1,587 without a notice, 232 with a notice). Scores for pharmacies without a notice were obtained by leave-one-out cross-validation.

**Supplementary Table S3.**
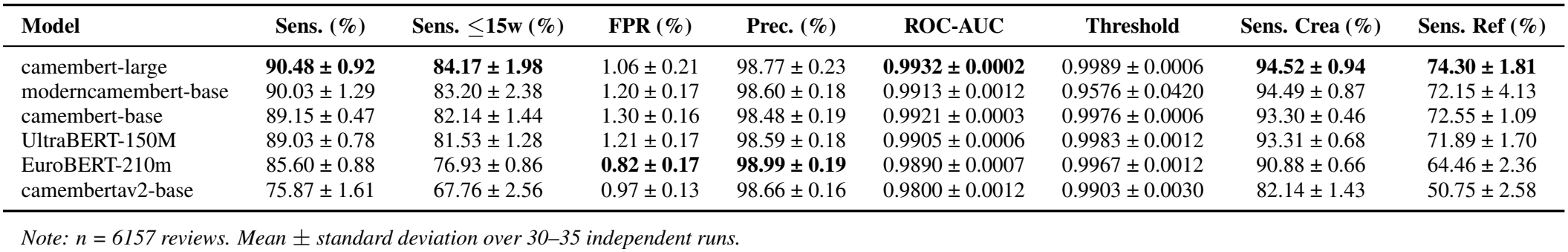
Performance comparison of the AI-detection models on the test set (n = 6157).

**Supplementary Fig. S3.**
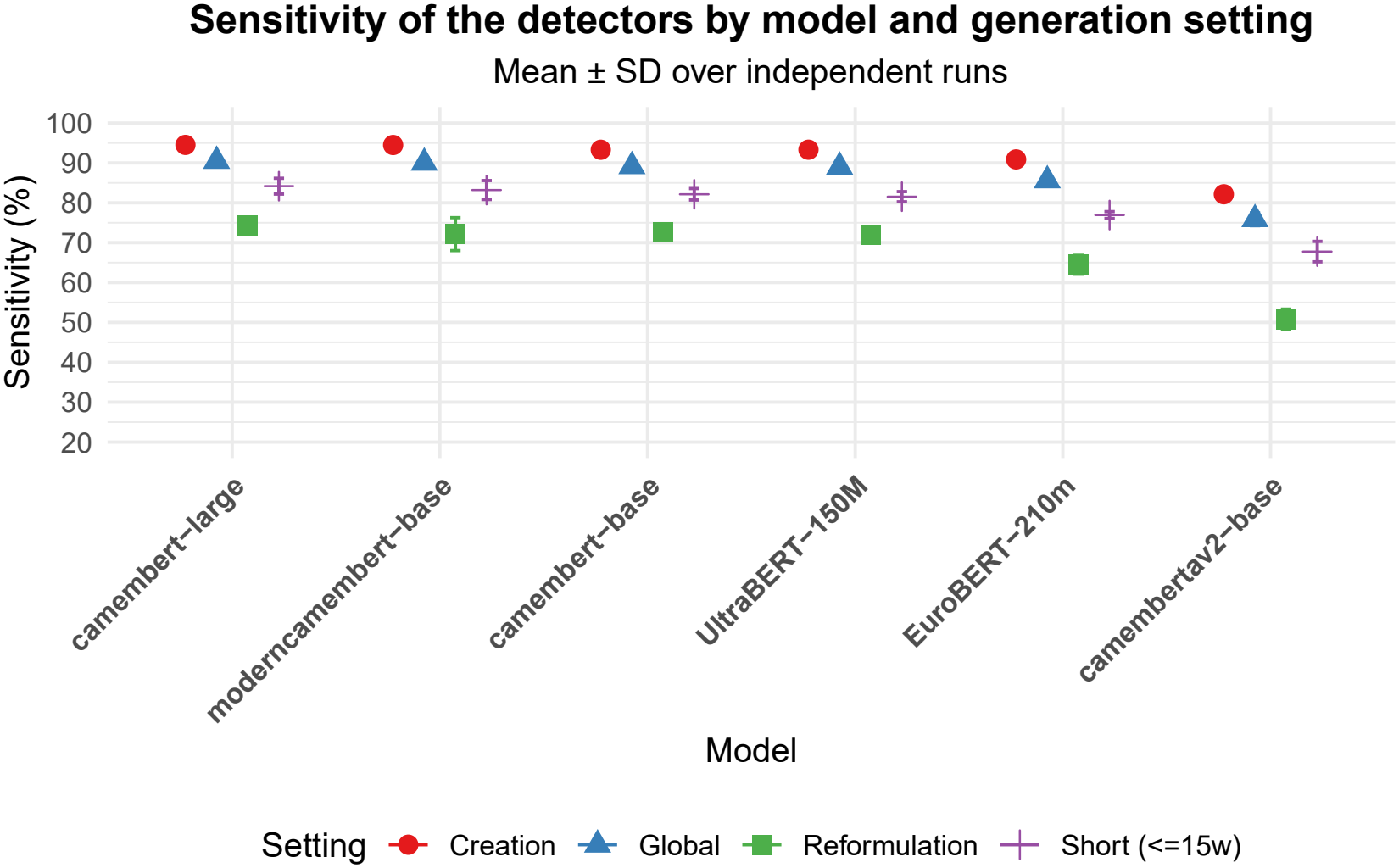
Comparison of detection sensitivity by model and generation mode. Points indicate the mean percentage and error bars represent the standard deviation over multiple independent runs.

## Notes

### Competing Interest Statement

The authors have declared no competing interest.

### Summary of Updates

Typographical errors have been corrected. Some elements that remained in French in the previous version have been translated into English.

## Bibliography

1. Y Alicia Hong, Chen Liang, Tiffany A Radcliff, Lisa T Wigfall, and Richard L Street. What Do Patients Say About Doctors Online? A Systematic Review of Studies on Patient Online Reviews. 21(4):e12521. ISSN 1438-8871. doi: 10.2196/12521.

2. Martin Emmert, Florian Meier, Frank Pisch, and Uwe Sander. Physician Choice Making and Characteristics Associated With Using Physician-Rating Websites: Cross-Sectional Study. 15(8):e187. ISSN 14388871. doi: 10.2196/jmir.2702.

3. Wei Lu and Hong Wu. How Online Reviews and Services Affect Physician Outpatient Visits: Content Analysis of Evidence From Two Online Health Care Communities. 7(4):e16185. ISSN 2291-9694. doi: 10.2196/16185.

4. Xi Han, Yongxi Lin, Wenting Han, Ke Liao, and Kefu Mei. Effect of Negative Online Reviews and Physician Responses on Health Consumers’ Choice: Experimental Study. 26:e46713. ISSN 1438-8871. doi: 10.2196/46713.

5. Doaa Al-Turkey, Tatiana Grieshofer, and Rasheed Mohammad. Online Patient Feedback About Medical Services: A Systematic Critical Review. 16(11):211. ISSN 2039-7283. doi: 10.3390/clinpract16110211.

6. Ordre National des Pharmaciens. Les pharmaciens - Panorama au 1er janvier 2026.

7. Yahya Ali Laghbi and Mohammed Al Dhoayan. Examining how customers perceive community pharmacies based on Google maps reviews: Multivariable and sentiment analysis. 15: 100498. ISSN 26672766. doi: 10.1016/j.rcsop.2024.100498.

8. Priya Bains, Fong Chan, James P. McCormack, and Aaron Sha. British Columbia community pharmacy during COVID-19: Describing the patient experience via Google reviews. 158(1):24–28. ISSN 1715-1635, 1913-701X. doi: 10.1177/17151635241281749.

9. Jingfang Liu, Yingyi Zhou, Xiaoyan Jiang, and Wei Zhang. Consumers’ satisfaction factors mining and sentiment analysis of B2C online pharmacy reviews. 20(1):194. ISSN 1472-6947. doi: 10.1186/s12911-020-01214-x.

10. Andrew M. Placona and Cheryl Rathert. Are Online Patient Reviews Associated With Health Care Outcomes? A Systematic Review of the Literature. 79(1):3–16. ISSN 1077-5587, 1552-6801. doi: 10.1177/10775587211014534.

11. Sonja Bidmon, Ossama Elshiewy, Ralf Terlutter, and Yasemin Boztug. What Patients Value in Physicians: Analyzing Drivers of Patient Satisfaction Using Physician-Rating Website Data. 22(2):e13830. ISSN 1438-8871. doi: 10.2196/13830.

12. Salma Patel, Rebecca Cain, Kevin Neailey, and Lucy Hooberman. General Practitioners’ Concerns About Online Patient Feedback: Findings From a Descriptive Exploratory Qualitative Study in England. 17(12):e276. ISSN 1438-8871. doi: 10.2196/jmir.4989.

13. Samir K. Trehan and Aaron Daluiski. Online Patient Ratings: Why They Matter and What They Mean. 41(2):316–319. ISSN 03635023. doi: 10.1016/j.jhsa.2015.04.018.

14. Andrew Yang, Fatima Rodriguez, and Jennifer P. Woo. Sex, Age, and Patient Experience in Cardiologist Reviews. 3(7):101046. ISSN 2772963X. doi: 10.1016/j.jacadv.2024.101046.

15. Baptiste Derombise, Cyril Desbarats, Olivia Guerin, Rémi Gaury, and Olivier Trost. Rating of French maxillofacial surgeons on their Google® business account: Current status and reflections for equitable communication. 125(4):101753. ISSN 24687855. doi: 10.1016/j.jormas.2023.101753.

16. Megan H. Trager, Dawn Queen, Weijia Fan, and Faramarz H. Samie. Patient Satisfaction of General Dermatologists: A Quantitative and Qualitative Analysis of 38,008 Online Reviews by Gender and Years of Experience. 2(2):100089. ISSN 26670267. doi: 10.1016/j.xjidi.2021.100089.

17. Elise Martin, Clifford Lober, Jeff Benabio, and Steven R Feldman. Addressing public criticism: A potential HIPAA violation. 24(6),. ISSN 1087-2108. doi: 10.5070/D3246040710.

18. France. Décret n°2026-156 du 3 mars 2026 modifiant le code de déontologie des pharmaciens et d’autres dispositions du code de la santé publique.

19. Gemma Team. Gemma 4 technical report. Technical report, Google DeepMind, 2026.

20. Chris Stokel-Walker. AI bot ChatGPT writes smart essays — should professors worry? pages d41586–022–04397–7. ISSN 0028-0836, 1476-4687. doi: 10.1038/d41586-022-04397-7.

21. Feng He. Catch the Patient, Not the AI: Collective Sensemaking in an Online Health Community.

22. Rachel C. Hill, Jane M. Grant-Kels, and Shari R. Lipner. Ethical considerations in responding to negative online reviews. 93(1):323–324. ISSN 01909622. doi: 10.1016/j.jaad.2024.05.026.

23. Shravika Mittal, Erin Kasson, Layna Paraboschi, Eleanor Laufenberg, Jiawei Zhou, Patricia A. Cavazos-Rehg, Tanushree Mitra, and Munmun De Choudhury. Follow the Rules (or Not): Community Norms and AI-Generated Support in Online Health Communities.

24. Louis Martin, Benjamin Muller, Pedro Javier Ortiz Suárez, Yoann Dupont, Laurent Romary, Éric De La Clergerie, Djamé Seddah and Benoît Sagot. CamemBERT: A Tasty French Language Model. In Proceedings of the 58th Annual Meeting of the Association for Computational Linguistics, pages 7203–7219. Association for Computational Linguistics,. doi: 10.18653/v1/2020.acl-main.645.

25. Wissam Antoun, Francis Kulumba, Rian Touchent, prefix=de la useprefix=true family=Clergerie, given=Éric, Benoît Sagot, and Djamé Seddah. CamemBERT 2.0: A Smarter French Language Model Aged to Perfection,.

26. Wissam Antoun, Benoît Sagot, and Djamé Seddah. ModernBERT or DeBERTaV3? Examining Architecture and Data Influence on Transformer Encoder Models Performance. In Proceedings of the 14th International Joint Conference on Natural Language Processing and the 4th Conference of the Asia-Pacific Chapter of the Association for Computational Linguistics, pages 3061–3074. The Asian Federation of Natural Language Processing and The Association for Computational Linguistics,. doi: 10.18653/v1/2025.ijcnlp-long.164.

27. Nicolas Boizard, Hippolyte Gisserot-Boukhlef, Duarte M. Alves, André Martins Ayoub Hammal, Caio Corro, Céline Hudelot, Emmanuel Malherbe, Etienne Malaboeuf, Fanny Jourdan, Gabriel Hautreux, João Alves, Kevin El Haddad, Manuel Faysse, Maxime Peyrard, Nuno M. Guerreiro, Patrick Fernandes, Ricardo Rei, and Pierre Colombo. EuroBERT: Scaling Multilingual Encoders for European Languages.

28. Jungwoo Park, Taewhoo Lee, and Jaewoo Kang. UltraBERT: Beyond the Limit of BERT Pre-Training.

